# Normal is All You Need: A Symmetry-Informed Inverse Learning Foundation Model for Neuroimaging Diagnostics

**DOI:** 10.64898/2026.04.10.26350553

**Authors:** Lola Siqi Wang, Cyrus Ayubcha, Yining Hua, Andrew Beam

## Abstract

**Background:** Developing generalizable neuroimaging models is often hindered by limited labeled data which has led to an increased interest in unsupervised inverse learning. Existing approaches often neglect geometric principles and struggle with diverse pathologies. We propose a symmetry-informed inverse learning foundation model to address these shortcomings for robust and efficient anomaly detection in brain MRI.

**Methods:** Our framework employs a reconstruction-to-embedding pipeline, trained exclusively on healthy brain MRI slices. A 2D U-Net uses a novel, symmetry-aware masking strategy to reconstruct a disorder-free slice. Difference maps are embedded into a 1024-dimensional latent space via a Beta-VAE. Anomaly scoring is performed using Mahalanobis distance. We evaluated generalization by fine-tuning on external lesion datasets, BraTS Africa (SSA), and the ADNI-derived Alzheimer’s disease cohort (Alz).

**Results:** On the source metastasis (Mets) dataset, the framework achieved high performance (AB1+MSE: 99.28% accuracy, 99.79% sensitivity). Generalization to the external lesion dataset (SSA) was robust, with the Symmetry ROC configuration achieving 91.93% accuracy. Transfer to the Alzheimer’s dataset (Alz) was more challenging, achieving a peak accuracy of 70.54% with a high false-positive rate, suggesting difficulty in separating subtle, diffuse changes.

**Conclusion:** The symmetry-informed inverse learning framework establishes a robust foundation model for neuroimaging, showing strong performance for focal lesions and successful generalization under domain shift. Limitations in diffuse neurodegeneration underscore the necessity for richer representations and multimodal integration to improve future foundation models.

**Summary Statement:** A symmetry-informed inverse learning framework trained on normal brain MRI achieved high accuracy for detecting focal lesions and demonstrated strong generalization across external datasets under domain shift.

**Key Points:**

● A symmetry-informed disorder-free reconstruction framework trained only on normal brain MRI achieved 99.28% accuracy and 99.79% sensitivity for metastasis detection on the BrainMetShare dataset, demonstrating non-inferior performance compared with all but one strategy while offering improved computational efficiency.
● The model generalized effectively to an external tumor dataset (BraTS SSA), achieving up to 91.93% accuracy using receiver operating characteristic–optimized thresholding with minimal fine-tuning.
● Embedding-based anomaly detection using Mahalanobis distance enabled consistent separation between normal and abnormal slices, supporting robust and interpretable anomaly detection across datasets.

## Introduction

Magnetic Resonance Imaging (MRI) remains the gold standard for most clinical neuroimaging purposes given its superior resolution and lack of ionizing radiation. Some of the earliest machine learning models were intended to facilitate diagnosis with neuroimaging data with notable success noted in their particular niche application. [1–3] Nevertheless, the complexity and variance in the radiological manifestation of neurological pathophysiology has led to many challenging problems in developing comprehensive and generalizable models for diagnosis. [1–3]

In contrast, unsupervised and self-supervised techniques have emerged as promising solutions overcoming these obstacles. [1–3] One approach has been the use of autoencoders, where poor reconstructions have been used as a means of identifying anomalous data. [4–6] For example, different autoencoder architectures were used to identify latent representation of normal brain structures and segment abnormal lesions in an unsupervised manner. [7] Further work, such as StRegA, has achieved better regularization in the latent space better capturing both local and global contexts.[8] More recently, He et al. explored an inverse learning approach in which models are trained exclusively on healthy brain scans, leveraging autoencoders to detect deviations from the normal training data used to develop the model [9–11, 25]

Existing methods have largely remained agnostic to geometric principles that could be leveraged to improve training efficiency in high-dimensional datasets. [12–14] Recent approaches that rely on training with healthy control populations have led to gains in lesion detection, but they continue to struggle when confronted with broader-spectrum diseases. [9, 15–16] In addition, the quantification and classification algorithms used in prior pipelines have been relatively rudimentary and limited in their effectiveness. Finally, the foundational use of these models has not been systematically explored in the context of fine-tuning to heterogeneous external datasets or to non-lesion pathologies such as Alzheimer’s disease (AD). [17–19] Collectively, these shortcomings underscore the need for adaptable foundation models capable of robustly and efficiently detecting both localized and widespread brain abnormalities. [20–24]

*Our contributions are as follows:*

● **Disorder-free self-supervised reconstruction model**: A reconstruction framework trained solely on healthy brain images, incorporating a symmetric masking strategy that reduces data complexity and improves efficiency while preserving anatomical fidelity to enhance anomaly detection.
● **Beta-VAE–based embedding extraction**: A Beta-VAE applied to difference images to learn informative embeddings that emphasize subtle deviations from normality for more sensitive anomaly detection.
● **Efficient anomaly classification in latent space**: A high-dimensional hypercube strategy for anomaly classification that scales effectively with latent dimensionality and provides transparent, interpretable thresholds to support diagnostic confidence.
● **Generalization to external datasets**: Demonstrated robust generalization across external lesion datasets and non-lesion pathologies.

## Methods

### Study design and overview

This study proposes and evaluates a symmetry-informed inverse-learning framework for anomaly detection in brain MRI. The approach learns a disorder-free reconstruction model from normal-appearing scans and quantifies abnormalities as deviations from the learned normative representation. The foundation reconstruction model was trained on the BrainMetShare dataset (Mets) using only normal axial slices. [26] Calibration for the classification threshold was performed in a held-out set. To investigate domain shift and model adaptability, the framework was further assessed on two external datasets: BraTS Africa (SSA) and an ADNI-derived Alzheimer’s disease cohort (Alz). [27, 28] For each external target domain, the reconstruction model was fine-tuned using normal slices within the respective dataset, and anomaly detection was evaluated using the same embedding-based inference pipeline. **Figure 1**

**Figure 1.**
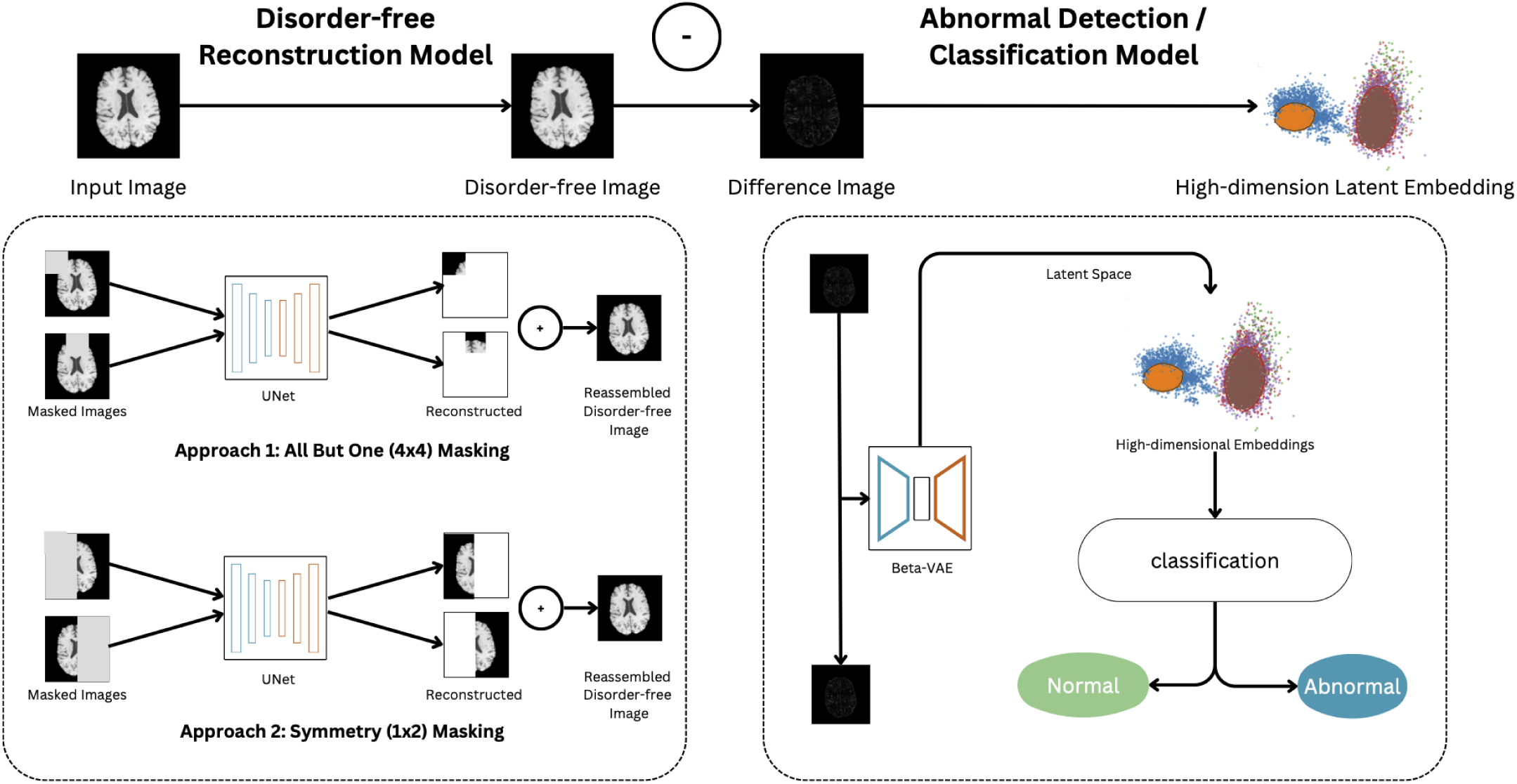
Reconstruction-to-embedding anomaly detection pipeline: Axial brain MRI slices are converted into self-supervised reconstruction tasks using two masking strategies: All-But-One (AB1; 4×4 grid masking; 16 pairs/slice) and Symmetry (left–right hemisphere prediction; 2 pairs/slice). A 2D U-Net reconstructs the masked target region, and reconstructed regions are reassembled into a predicted disorder-free slice. A difference image is computed (original-reconstruction) and embedded by a Beta-VAE into a 1024-dimensional latent embedding. Anomalies are scored by Mahalanobis distance in embedding space using percentile thresholds (internal dataset) and ROC-optimized thresholds (external dataset).

### Data sources and cohort construction

Datasets and split statistics are summarized in **Table 1**. We used BrainMetShare to train the foundation reconstruction model and to perform internal validation; we assessed external domain shifts with fine-tuning on SSA and Alz.

**Table 1.**
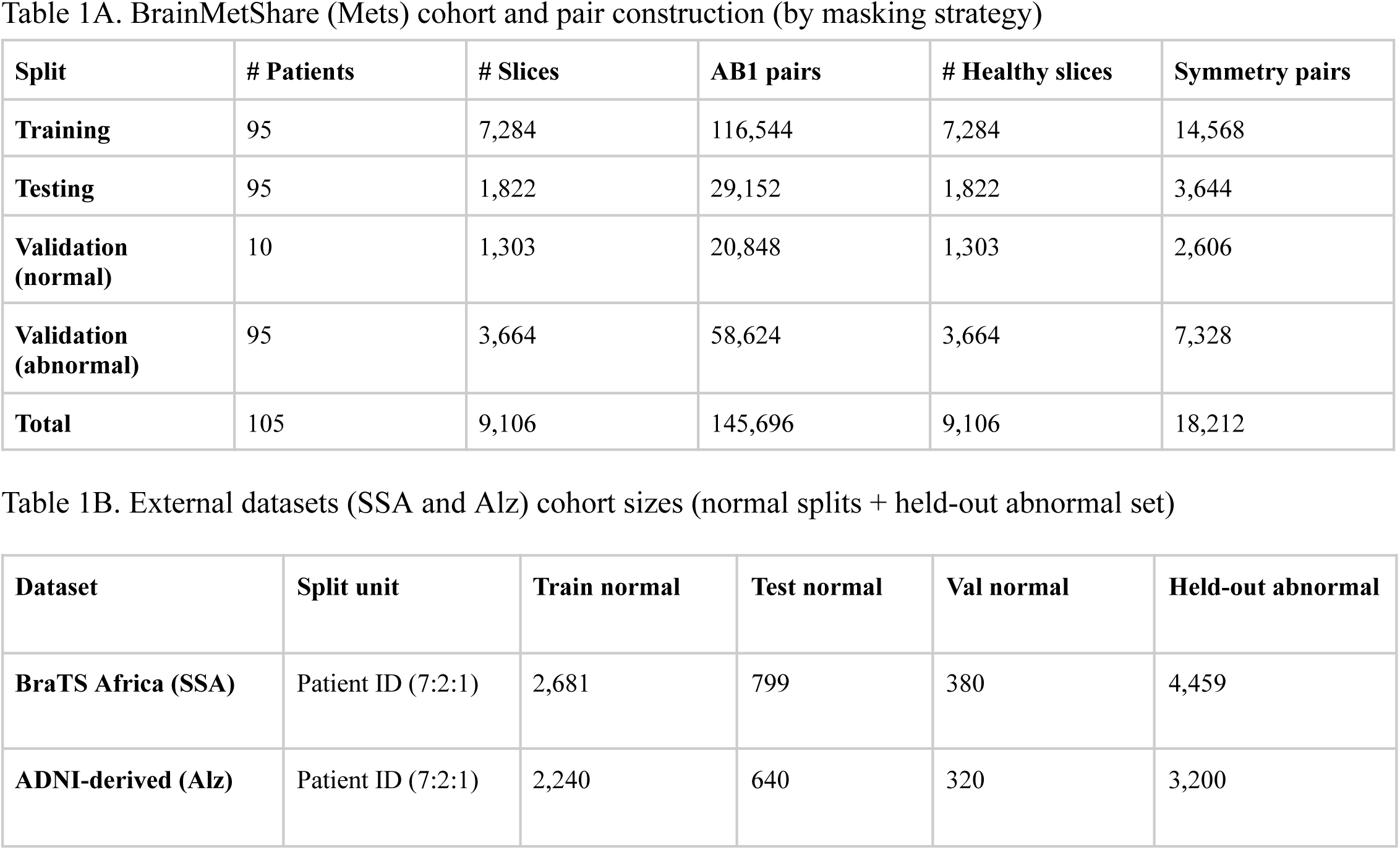
Datasets and partitioning used for pretraining and external validation/fine-tuning (updated to match slides)

*BrainMetShare* is a publicly available, expert-curated brain MRI dataset of patients with cerebral metastases. We used the T1-weighted pre-contrast sequence and constructed a subcohort to support disorder-free reconstruction training, i.e., including lesion-containing slices in training would bias reconstruction toward pathology and reduce contrast at inference. Cohort construction followed two steps (**Fig. 2**). First, the 10 metastasis-negative patients were selected to form a validation-normal cohort. Second, among 95 metastasis-positive patients, we extracted normal slices to build the reconstruction corpus and reserved all abnormal slices as a held-out abnormal evaluation set. Normal slices from the 95 metastasis-positive patients were split at the image (axial slice) level into training (80%) and testing (20%), while abnormal slices were placed entirely into the validation-abnormal set. This yielded 7,284 training healthy slices, 1,822 testing healthy slices, 1,303 validation-normal healthy slices (from 10 metastasis-negative patients), and 3,664 validation-abnormal slices. External adaptation experiments were performed on *SSA*. Patients were split into train:test:validation sets (ratio 7:2:1). The train/test/validation partitions were formed from normal slices only, while abnormal slices were held out for anomaly evaluation. *SSA* included 2,681 / 799 / 380 normal slices in train/test/validation (42 / 12 / 6 patients) and 4,459 abnormal slices in the held-out abnormal set. We also evaluated transfer to a diffuse neurodegenerative condition using an *Alz* with patient-level splitting (7:2:1), using normal slices for train/test/validation and a held-out abnormal set for evaluation. The *Alz* partition comprised 2,240 / 640 / 320 normal slices in train/test/validation and 3,200 abnormal slices.

**Figure 2.**
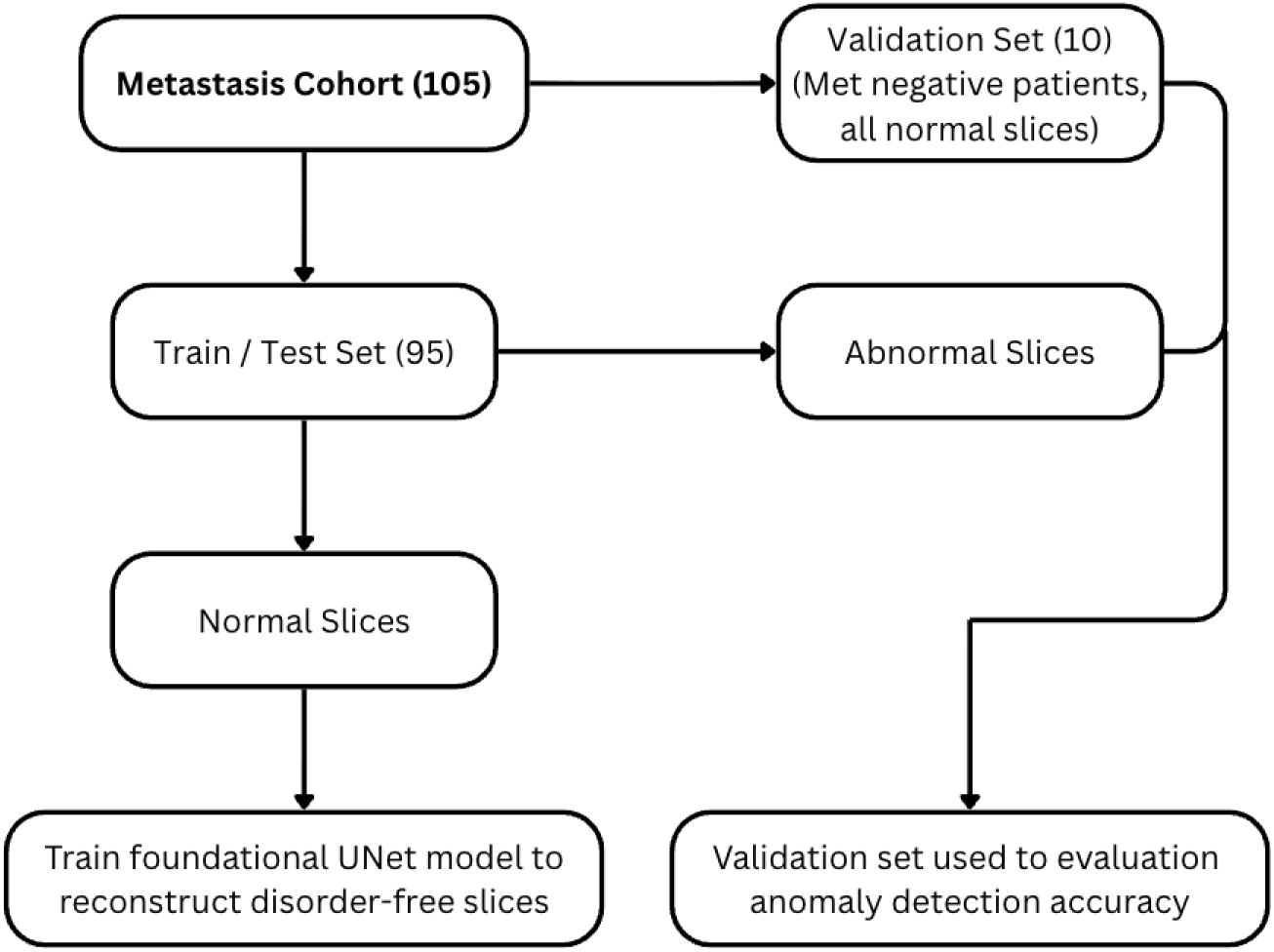
Cohort Partition and Data Flow for UNet Training.

### Image Preprocessing and Representation Construction

All volumes were converted into 2D axial slices and resized to 256×256. We applied a standardized preprocessing pipeline that (i) crops slices so the brain is centered with consistent margins, (ii) computes an edge map per slice, and (iii) constructs a binary diluted contour (ROI) mask representing brain tissue used for ROI-restricted loss computation. The edge map is used as the second input channel, while the ROI mask is used only for loss/evaluation masking. Intensity slices and edge maps were normalized prior to model input. Further implementation details are provided in **Appendix S1**.

### Self-supervised Masking Strategies

We generated self-supervised image reconstructions from each 2D axial slice (256×256) using two masking strategies. In the All-But-One (AB1) strategy, each slice was partitioned into a 4×4 grid and one patch was masked as the reconstruction target while the remaining patches provided context, yielding 16 masked pairs per slice. In the Symmetry strategy, each slice was divided into left and right hemispheres; one hemisphere served as input and the contralateral hemisphere served as the reconstruction target. Image-mask pairs were excluded when the target region contained no brain signal after preprocessing to avoid training on a non-informative background. For both strategies, the model input was a two-channel tensor formed by concatenating the masked image with its corresponding edge-map channel. Standard 2D augmentations (including shear and horizontal flip) were applied to masked pairs.

### Reconstruction Model and Training

We trained a 2D U-Net reconstructor under each masking strategy using two loss functions, resulting in four model variants (AB1×{MSE, ROI loss} and Symmetry×{MSE, ROI loss}). Mean squared error (MSE) was computed over pixels in the reconstruction target. ROI loss was defined as an MSE restricted to brain tissue using the binary diluted contour mask m_i_∈{0,1}:

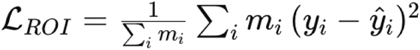

Here, *y_i_* denotes the ground-truth pixel intensity at pixel *i* 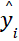 is the corresponding reconstructed pixel value predicted by the U-Net, and *m_i_* is the binary brain mask indicating whether pixel *i* lies inside brain tissue (*m_i_* = 1) or outside (*m_i_i* = 0). The summation is taken over all pixels in the image, and normalization by 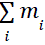 ensures that the loss is averaged only over brain regions, excluding background and non-anatomical areas.

For fair comparison across loss types, validation forward-pass loss was computed within the brain ROI and empty/background-only images were excluded, consistent with the slide definition. Each model was trained for up to 300 epochs with best-checkpoint selection based on the lowest validation forward-pass loss.

### Difference Representation and Beta-VAE Embeddings

After U-Net training, each slice was reconstructed to generate a predicted disorder-free image, and a difference map was computed between the original slice and its reconstruction. We then trained a Beta-VAE on difference maps to produce a 1024-dimensional latent embedding per slice for downstream anomaly detection. A separate Beta-VAE was trained for each reconstruction configuration (masking strategy × loss). Architecture and hyperparameters are provided in Appendix S1.

### Anomaly Detection in Embedding Space

For each slice, we computed a 1024-dimensional latent embedding z using the trained Beta-VAE. We then defined an anomaly score as the Mahalanobis distance from test sample (z) to a multivariate normal reference distribution estimated from embeddings of normal slices. Specifically, let μ\muμ and Σ\SigmaΣ denote the empirical mean vector and covariance matrix of the reference normal embeddings; the anomaly score for embedding z is

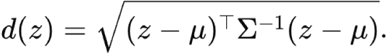

When Σ was ill-conditioned, we used a Moore-Penrose pseudoinverse in place of Σ^-1^ for numerical stability.

For Mets, we report two evaluation experiments to discern optimal thresholding value: (i) thresholding evaluation in validation-normal percentile, and (ii) 50% of validation-normal sensitivity analysis in which half of the validation-normal embeddings define the reference distribution and the remaining half, together with all abnormal slices, is used for evaluation.

For SSA and Alz, we performed reconstruction fine-tuning (epochs = 50) by using the pretrained U-Net; we varied the amount of available normal training data (0%, 25%, 50%, 100%) (**Fig. 3**). Beta-VAE weights were kept fixed and embeddings were computed using the pretrained Beta-VAE checkpoints. For evaluation, normal embeddings were split into two halves: the first half defined the Mahalanobis reference distribution, while the second half was evaluated alongside held-out abnormal slices. Classification thresholds were determined by ROC analysis (maximizing TPR - FPR) on the evaluation set of distances, and accuracy, sensitivity, and specificity were reported at that ROC-derived threshold. Sensitivity analyses evaluating the effect of normal reference-set size on embedding-space distance estimation, and additional fine-tuning reconstruction-loss diagnostics, are reported in Appendix S2.

**Figure 3.**
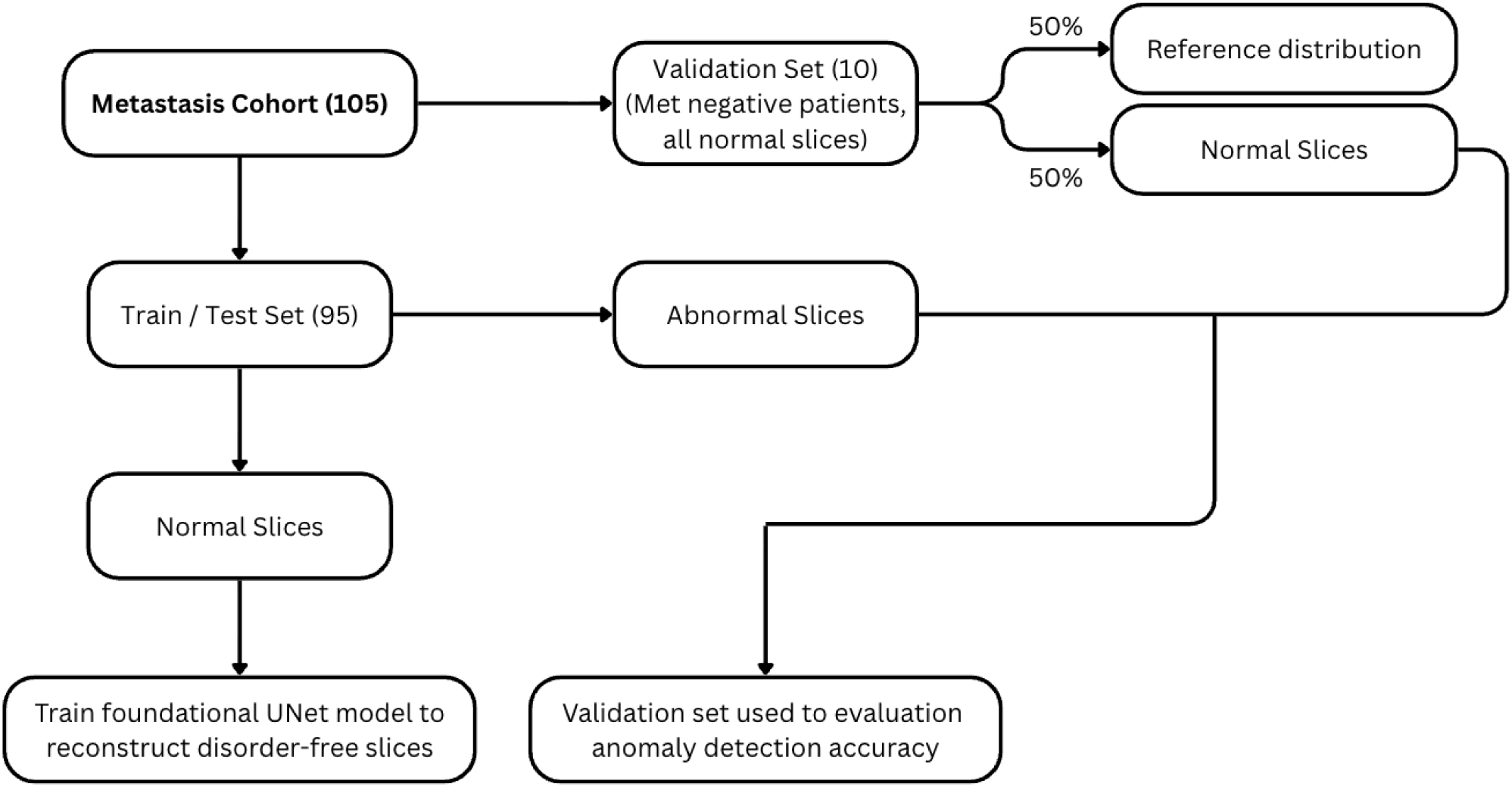
Validation Set Construction for Anomaly Detection Framework.

## Results

### Disorder-Free Reconstruction Performance (BrainMetShare)

All four U-Net configurations trained on the normal-only BrainMetShare (Mets) corpus converged with relatively low reconstruction error, indicating stable disorder-free reconstruction across models. The AB1 architecture with ROI loss achieved the best overall performance, with the lowest training loss (1.13×10⁻⁵), test loss (4.16×10⁻⁵), and forward-pass validation loss (6.009×10⁻⁴), despite having the same number of parameters as the other configurations (23.6M) and being selected at epoch 66 out of a 300-epoch training budget. **Table 2**

**Table 2.** U-Net reconstruction performance on normal-only corpus (Mets).“Epochs (max/best)” denotes the maximum training budget and the selected best checkpoint.

|  | All But One |  | Symmetry |  |
| --- | --- | --- | --- | --- |
|  | MSE | ROI loss | MSE | ROI loss |
| Params | 23,602,177 | 23,602,177 | 23,602,177 | 23,602,177 |
| Total / Best Epoch | 300 / 50 | 300 / 66 | 300 / 57 | 300 / 121 |
| Training Loss | 6.45e-05 | 1.13e-05 | 2.40e-04 | 1.18e-04 |
| Testing Loss | 6.58e-05 | 4.16e-05 | 4.44e-04 | 4.05e-04 |
| Validation Loss | 6.494e-04 | 6.009e-04 | 7.006e-04 | 7.632e-04 |

### Metastasis anomaly detection: Embedding-space thresholding

The validation set consists of 1,303 normal slices (26.23%) and 3,664 abnormal slices (73.77%), used to evaluate the anomaly detection performance of the Beta-VAE embeddings. **Fig. 4** presents a two-dimensional visualization of the learned latent space, obtained by projecting the high-dimensional embeddings onto the first two principal components using PCA.

**Figure 4.**
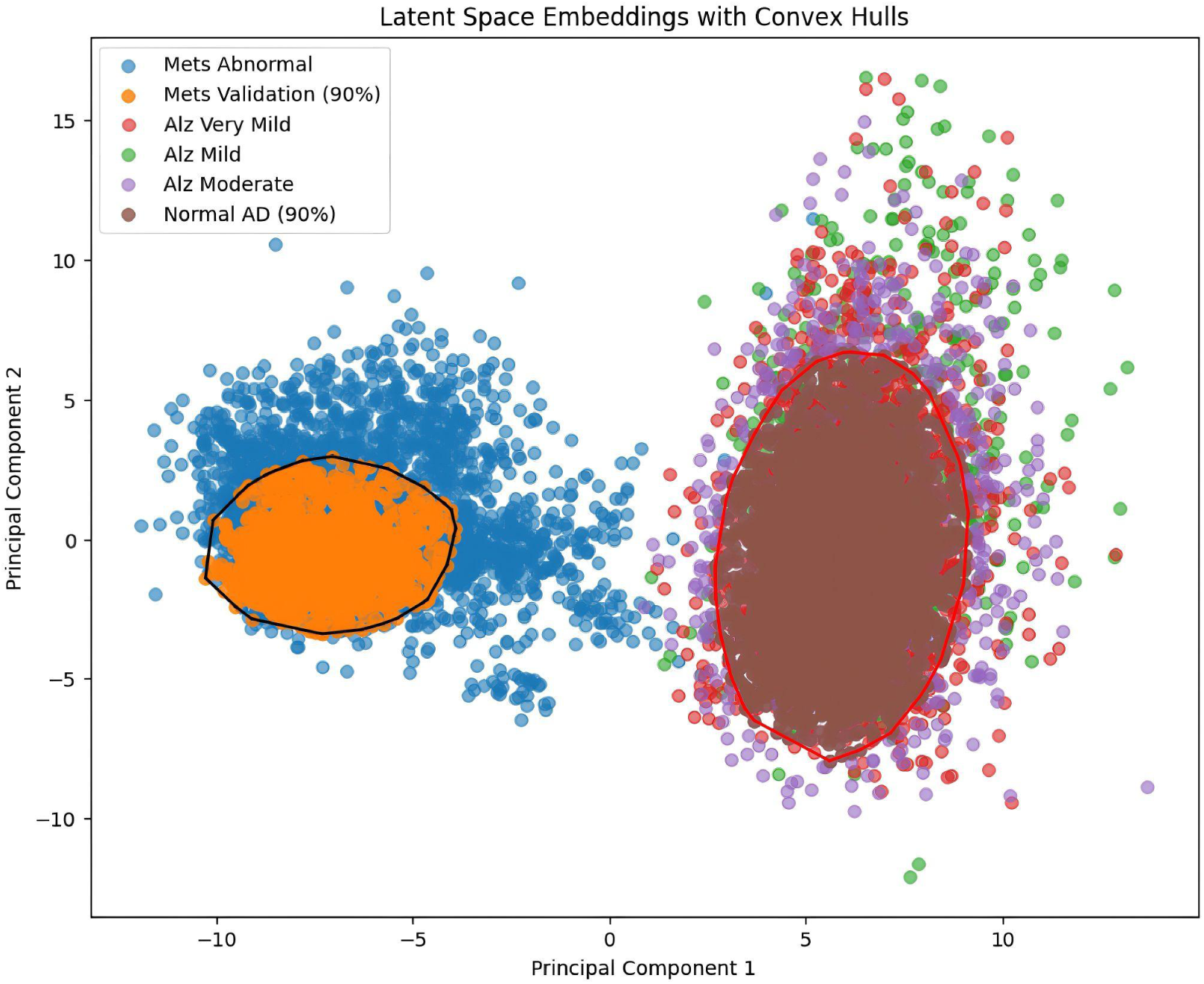
Latent Space Embeddings of Brain MRI Cohorts with Convex Hulls. This visualizes the 2D latent space representation of brain MRI embeddings using Principal Component Analysis (PCA) for dimensionality reduction. Different clinical cohorts are color-coded, and convex hulls are overlaid to illustrate the spatial distribution and separability of normal and abnormal populations in the embedding space. A clear boundary is shown in the latent space that separates the normal and abnormal slices, highlighting the model’s effectiveness in learning meaningful representations for distinguishing pathology.

Percentile-based discrimination was employed using the Mahalanobis distance in the 1024-dimensional Beta-VAE embedding space where the respective thresholds were derived from a reference distribution (i.e. validation-normal embeddings) to determine the optimal calibration of the operating point (Table 3A). At the 98th-percentile operating point, AB1+MSE achieved 99.28% accuracy with 99.79% sensitivity, and symmetry masking produced comparable performance. At more stringent operating points, sensitivity and accuracy decreased as expected (e.g., AB1+MSE sensitivity 97.76% and accuracy 98.38% at the 100th percentile). A sensitivity analysis using half of the validation-normal embeddings to estimate the reference distribution is provided in **Appendix S2 (Table S1)**.

**Table 3.** BrainMetShare (Mets) anomaly detection: Comparison of reference-set sizes. Table 3A. Full Reference Set (Reference = Validation-Normal Distribution)

| Threshold | Metric | AB1 (MSE) | AB1 (ROI) | Sym (MSE) | Sym (ROI) |
| --- | --- | --- | --- | --- | --- |
| 95% | Accuracy | 98.56% | 96.96% | 97.93% | 97.33% |
| 95% | Sensitivity | 99.93% | 97.72% | 99.06% | 98.23% |
| 95% | Specificity | 94.95% | 94.95% | 94.95% | 94.95% |
| 98% | Accuracy | 99.28% | 96.03% | 98.09% | 97.21% |
| 98% | Sensitivity | 99.79% | 95.32% | 98.14% | 96.94% |
| 98% | Specificity | 97.94% | 97.94% | 97.94% | 97.94% |
| 99% | Accuracy | 99.01% | 95.04% | 97.78% | 96.05% |
| 99% | Sensitivity | 99.03% | 93.57% | 97.34% | 96.96% |
| 99% | Specificity | 98.94% | 98.94% | 98.94% | 98.94% |
| 100% | Accuracy | 98.38% | 91.32% | 94.15% | 92.98% |
| 100% | Sensitivity | 97.76% | 88.04% | 91.95% | 90.32% |
| 100% | Specificity | 100% | 100% | 100% | 100% |

We evaluated sensitivity to reference-set size by re-estimating the normal reference distribution parameters μ, Σ using 50% of the validation-normal embeddings and then evaluating anomaly detection on the remaining validation-normal embeddings together with all abnormal slices (**Table 3B**).

**Table 3B.** Reduced Reference Set (Reference = 50% of Validation-Normal)

| Threshold | Metric | AB1 (MSE) | AB1 (ROI) | Sym (MSE) | Sym (ROI) |
| --- | --- | --- | --- | --- | --- |
| 90% | Accuracy | 78.40% | 69.43% | 70.14% | 70.73% |
| 90% | Sensitivity | 79.94% | 69.14% | 69.92% | 70.67% |
| 90% | Specificity | 70.20% | 70.95% | 71.32% | 71.07% |
| 95% | Accuracy | 74.67% | 64.51% | 65.58% | 65.37% |
| 95% | Sensitivity | 74.20% | 61.86% | 63.35% | 62.57% |
| 95% | Specificity | 77.18% | 78.55% | 77.43% | 80.17% |
| 99% | Accuracy | 68.57% | 52.63% | 54.04% | 55.52% |
| 99% | Sensitivity | 65.28% | 53.81% | 47.53% | 49.41% |
| 99% | Specificity | 86.03% | 89.96% | 88.53% | 87.91% |

Performance decreased across all model variants (e.g., AB1+MSE accuracy 78.40% at the 90th percentile; 74.67% at the 95th; 68.57% at the 99th), indicating that mean/covariance estimation for high-dimensional Mahalanobis scoring benefits from larger normal reference sets.

### External domain shift: BraTS Africa (SSA)

We reconstructed disorder-free slices and quantified reconstruction quality using mean squared error. As shown in Table S2, baseline errors before fine-tuning were lower for the AB1 model (6.5e-04) than for the Symmetry model (7.0e-04), suggesting better generalization in the AB1 setting. Fine-tuning the models on increasing proportions of in-domain data (0%, 25%, 50%, 100%) led to progressive reductions in reconstruction error. For example, at 100% fine-tuning, AB1 achieved an MSE of 9.7e-04, while Symmetry reached 1.09e-03. These improvements in reconstruction quality form the basis for downstream anomaly detection using Mahalanobis distance, as described below. Under domain shift, ROC-optimized Mahalanobis thresholding consistently outperformed the KMeans baseline across all fine-tuning scales for SSA anomaly classification (**Fig. 5)**. The strongest result was obtained with Symmetry ROC at 25% fine-tuning, achieving 91.93% accuracy, 91.41% sensitivity, and 93.16% specificity. This substantially exceeded the best AB1 ROC configuration, which reached 84.78% accuracy at 100% fine-tuning.

**Figure 5.**
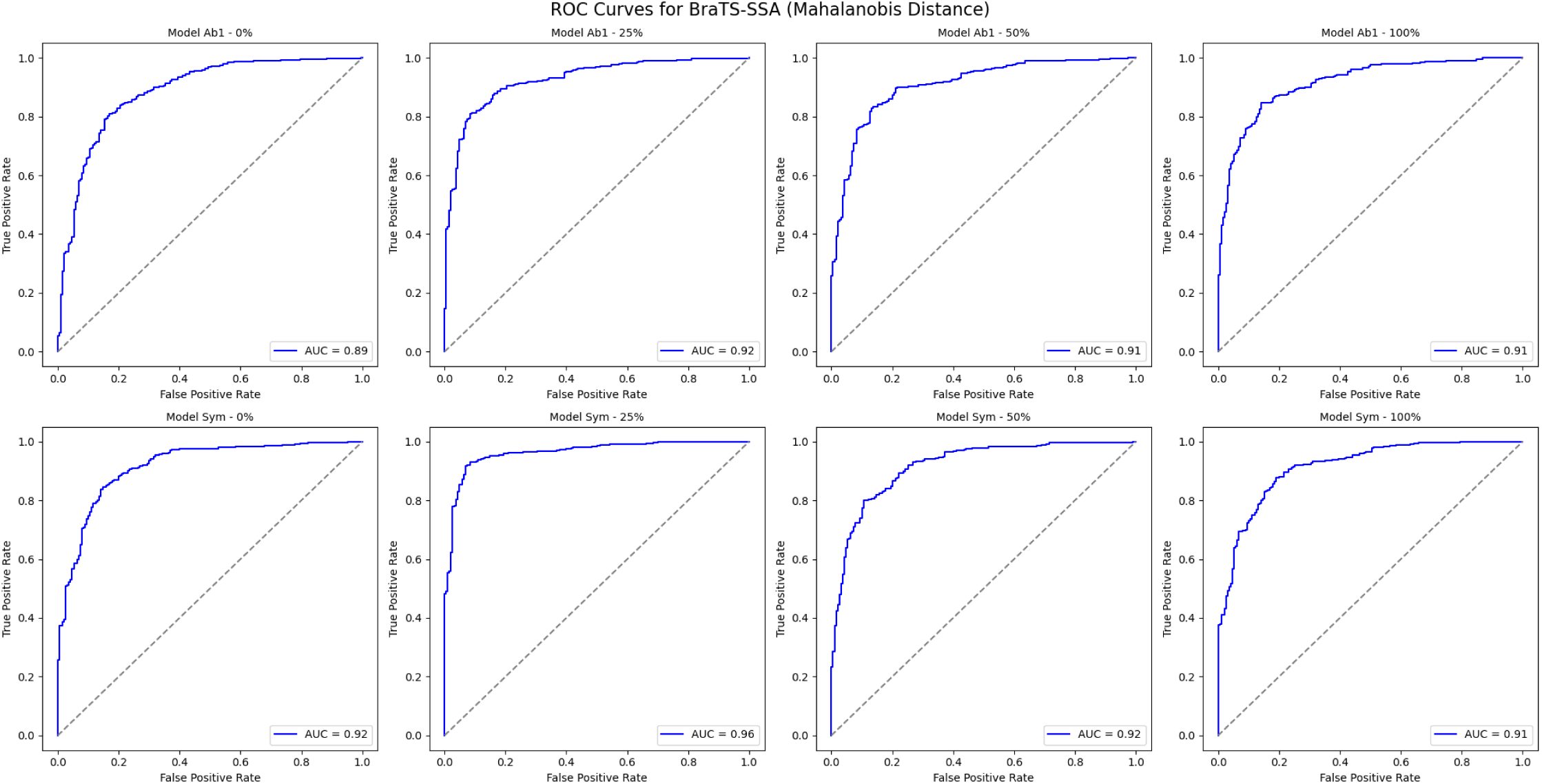
ROC Curves Across Different Proportion of Data Used for Fine-tuning BraTS-SSA Model. This figure presents the ROC (Receiver Operating Characteristic) curves comparing the anomaly detection performance of both Ab1 and Sym, across different fine tuning proportions (0%, 25%, 50%, and 100% of available data). Each curve illustrates the trade-off between true positive and false positive rates, with performance evaluated using Mahalanobis distance on Beta-VAE latent embeddings. The AUC (Area Under the Curve) values indicate consistently high discriminative power across settings, with the Sym model slightly outperforming Ab1 in most cases, especially at lower training percentages.

Symmetry ROC performance was not monotonic with respect to the amount of fine-tuning data: for instance, accuracy was 82.61% at 50% fine-tuning but 91.93% at 25%, indicating that the interaction between fine-tuning scale and the resulting embedding geometry can shift the optimal operating point for ROC thresholding. As a clustering-based baseline, KMeans (k = 2) applied to embeddings yielded consistently lower performance than ROC-optimized thresholding at all fine-tuning scales. The strongest KMeans result was Symmetry KMeans at 25%, with 82.37% accuracy, 74.01% sensitivity, and 92.37% specificity, which still fell short of the corresponding Symmetry ROC result at the same scale. Fine-tuning reconstruction-loss trajectories for each training scale are reported in Appendix S2 (Tables S2–S3).

**Table 4.** SSA anomaly classification across fine-tuning scales: ROC-optimized thresholding vs KMeans baseline.

| Method | Metric | Proportion of<br>BRATs Data:<br>0% | Proportion of<br>BRATs Data:<br>25% | Proportion of<br>BRATs Data:<br>50% | Proportion of<br>BRATs Data:<br>100% |
| --- | --- | --- | --- | --- | --- |
| AB1 ROC | Accuracy | 81.37% | 83.85% | 83.85% | 84.78% |
|  | Sensitivity | 80.62% | 80.62% | 82.60% | 84.36% |
|  | Specificity | 83.16% | 91.58% | 86.84% | 85.79% |
| Sym ROC | Accuracy | 84.63% | 91.93% | 82.61% | 85.56% |
|  | Sensitivity | 84.36% | 91.41% | 79.74% | 87.44% |
|  | Specificity | 85.26% | 93.16% | 89.47% | 81.05% |
| AB1 KMeans | Accuracy | 69.42% | 69.66% | 69.06% | 68.35% |
|  | Sensitivity | 66.74% | 65.64% | 67.18% | 64.10% |
|  | Specificity | 72.63% | 74.47% | 71.32% | 73.42% |
| Sym KMeans | Accuracy | 71.58% | 82.37% | 71.10% | 70.50% |
|  | Sensitivity | 67.18% | 74.01% | 66.30% | 69.38% |
|  | Specificity | 76.84% | 92.37% | 76.84% | 71.84% |

### External domain shift with fine-tuning: ADNI-derived Alzheimer’s Dataset

Transfer to diffuse neurodegeneration (AD) was more challenging than transfer to metastasis or SSA. Using ROC-optimized Mahalanobis thresholds, AB1 ROC achieved its best result at 100% fine-tuning, with 70.54% accuracy, 71.84% sensitivity, and 44.38% specificity (threshold 44%) (**Fig. 6**). This indicates reasonable detection of anomalies but with a high false-positive rate (Table S4). In contrast, Symmetry ROC tended to favor higher specificity at the cost of sensitivity. Its best accuracy was 51.07% at 25% fine-tuning, with 50.28% sensitivity, 66.88% specificity, and a threshold of 67%. At 0% fine-tuning, Symmetry ROC reached very high specificity (83.75%) but very low sensitivity (34.66%), further illustrating this bias. This pattern is consistent with a more subtle and spatially distributed signal in Alzheimer’s disease, where slice-level reconstruction residuals are less sharply separable than focal lesions, making it harder for both AB1 and symmetry-based embeddings to cleanly distinguish normal from abnormal slices under domain shift.

**Figure 6.**
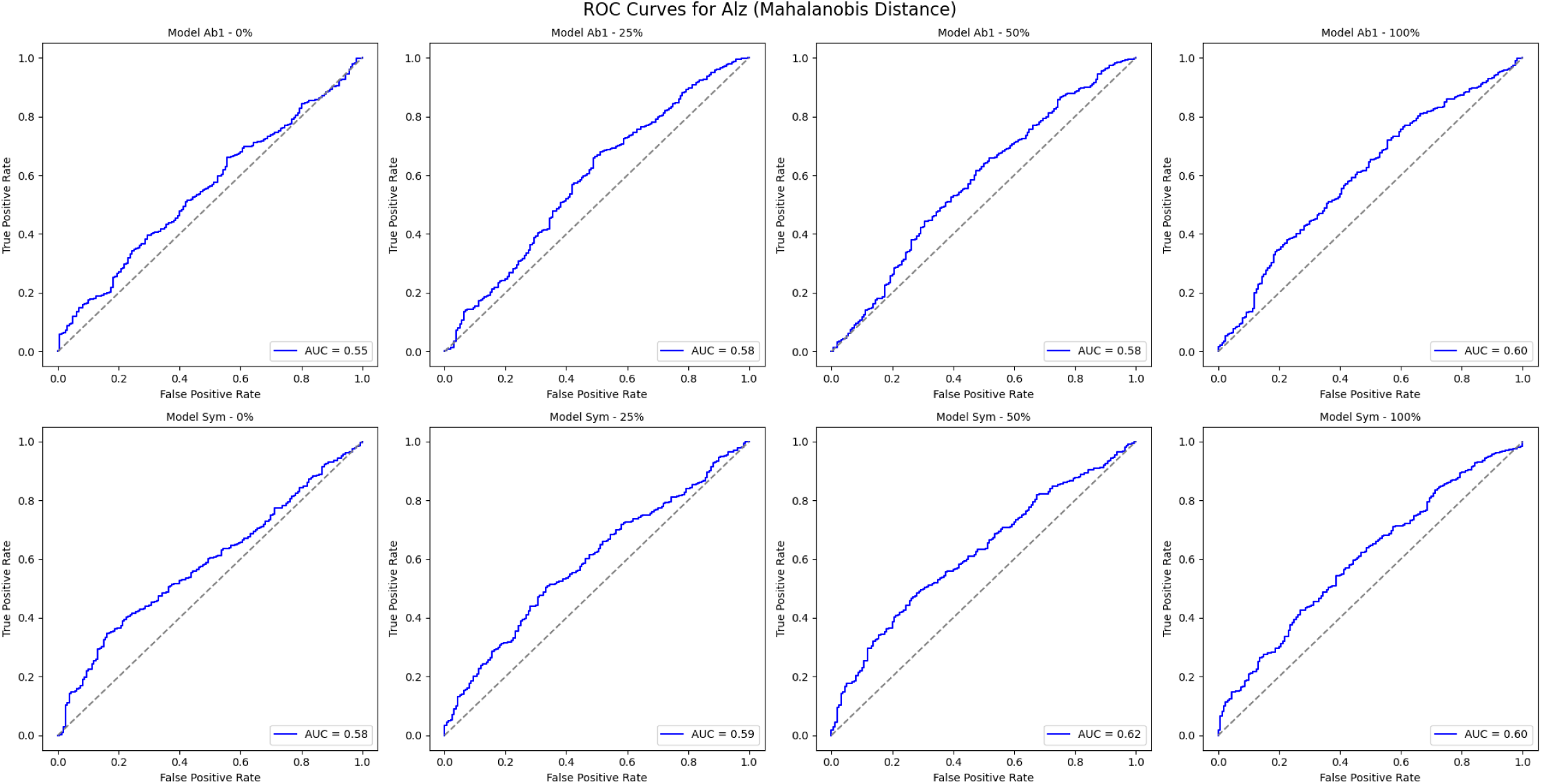
ROC Curves Across Different Proportion of Data Used for Fine-tuning BraTS-SSA Model.

## Discussion

This work presents a symmetry-informed inverse learning framework for anomaly detection in brain MRI, trained exclusively on disorder-free data and evaluated in both focal and diffuse brain pathologies [7–9,25,29]. By combining symmetry-aware self-supervised reconstruction, difference-map–based Beta-VAE embeddings, and Mahalanobis distance-based anomaly scoring, we show that a single foundation model can support robust lesion detection across domains, while also revealing important limitations in the context of diffuse neurodegeneration [1–9,12,25]. On the source metastasis dataset, all reconstruction configurations trained on normal-only slices converged to low reconstruction error, which supports the feasibility of learning a stable disorder-free representation without exposure to pathology [1–3,5–9,11,25]. While the combination of a patch-based masking strategy and an ROI-restricted loss provided the most favorable reconstruction behavior, symmetry-based methods were comparable [7–9,29].

Percentile-based thresholding on Mahalanobis distance in this embedding space produced strong anomaly discrimination [1–3,5–9,12]. Normal and metastatic slices were well separated, and performance remained high across a range of operating points that traded sensitivity for specificity. These findings indicate that focal lesions induce reconstruction residuals that are both consistent and distinctive relative to the normal manifold learned from healthy slices [1–3,5–9,11]. These results are consistent with prior work on inverse supervised learning, which has demonstrated that disorder-free frameworks can be non-inferior to, or outperform, conventional baseline classifiers for broad-spectrum anomaly detection tasks [10,11,16,19,25]. A sensitivity analysis that used fewer normal samples to estimate the reference distribution showed a clear degradation in anomaly detection. When only a limited set of normal examples is available for calibration, distance estimates can become unstable and lead to reduced performance [1,2,6–9,16].

Under domain shift to an external lesion dataset, the same embedding-based framework remained effective when combined with modest fine-tuning of the reconstruction model on target domain normal slices [9,13–16,18]. Across a range of fine-tuning scales, ROC-optimized Mahalanobis thresholding consistently outperformed a simple clustering baseline. This pattern held for both masking strategies, with symmetry-based masking showing robust transfer performance [7–9,15,29].

Symmetry masking introduces a neuroanatomically grounded prior by explicitly modeling left-right hemispheric correspondence [7–9,29]. In the external lesion setting, this prior appears to be especially helpful, likely because many lesions distort expected symmetry in a way that is shared across sites and scanners [7–9,25,29]. Interestingly, performance under symmetry masking did not improve monotonically with increasing amounts of fine-tuning data. Instead, it peaked at an intermediate fine-tuning scale. This non-monotonic behavior suggests that there is a balance between adapting the reconstructor to the target distribution and preserving the disorder-free structure learned from the source, reflecting known trade-offs with catastrophic forgetting during medical domain expansion [14,15,18,30]. Excessive adaptation may shift the reconstruction residuals in a way that weakens separation between normal and abnormal embeddings.

Transfer to diffuse neurodegeneration in an Alzheimer’s-related cohort proved more challenging than transfer to lesion-based datasets [9,10,16,19]. Even with fine-tuning on target domain normal slices, separation between normal and abnormal embeddings in AD was limited, and the best configurations achieved only moderate accuracy with a clear sensitivity-specificity trade-off [9,10,16,19]. Patch-based masking tended to detect more abnormal slices but at the cost of more false positives, whereas symmetry-based masking usually yielded higher specificity while missing more abnormalities [7–9,16,19]. This pattern is consistent with subtle, gradual, and spatially diffuse Alzheimer’s-related changes, which may not generate sharp reconstruction errors and can be partly absorbed as normal variation during training and adaptation, suggesting a possible need for a larger corpus of normal subjects [9,10,16,19].

This study has several limitations. The evaluation was restricted to a small number of datasets, focused mainly on a single MRI contrast and two broad classes of pathology, and relied on 2D slice-based models that do not exploit full 3D structure or multimodal information [7–9,13,17,18,20]. The embedding model was kept fixed during external fine-tuning, which simplifies the pipeline but may limit adaptation to new domains, and all analyses were conducted at the slice level rather than the patient level, which differs from typical clinical decision-making [14–16,18]. Future work should extend the framework to 3D or multi-view architectures, incorporate additional MRI sequences, jointly adapt both reconstructor and embedding models under domain shift, test across more diverse populations and disorders, and develop patient-level aggregation strategies that can be compared directly with radiologist performance [13–15,17,18,20–24].

The proposed symmetry-informed inverse learning framework shows how disorder-free training, anatomically motivated masking, and embedding-space anomaly detection can form the basis of a neuroimaging foundation model [7–9,21–24,25]. The approach performs strongly for focal lesions, generalizes to an external lesion dataset under domain shift, and provides transparent, tunable anomaly scores that can be aligned with clinical priorities [1–3,5–9,11,20,21–25]. At the same time, its more limited performance in diffuse neurodegeneration underscores the need for richer representations and multimodal integration, and points to clear avenues for improving future foundation models for brain disease [9,10,13,16–19,21–24,31].

## Data Availability

All data used in this study are publicly available. BrainMetShare data are available at https://doi.org/10.71718/z66c-qr59. BraTS datasets are available at https://www.med.upenn.edu/cbica/brats2023/ upon registration. ADNI data are available at http://adni.loni.usc.edu upon application and approval.

## Appendix S1. Implementation details (preprocessing, pair generation, model training)

### S1.1 Image preprocessing

Input format. All experiments use 2D axial slices resized/cropped to 256×256.

Skull stripping and brain mask. Volumes were skull stripped using FSL BET to remove non-brain tissue and to obtain a binary brain mask used downstream for ROI construction.

Cropping. Skull-stripped images were cropped to center the brain and standardize spatial margins across scans.

Edge-map channel. An edge map was computed per slice using a Canny-based procedure, producing a single-channel edge image aligned to the intensity slice. The edge map was concatenated with the masked slice to form a 2-channel input.

ROI mask for ROI-restricted loss. A dilated brain-contour mask was derived from the brain mask to constrain ROI loss computation to brain tissue (and downweight background/non-informative regions). The ROI mask is binary with values in {0,1}.

Normalization. Intensity slices and edge maps were z-score normalized prior to model input.

### S1.2 Self-supervised masking pair construction

We generated image–target pairs from each preprocessed 256×256 slice under two strategies.

All-But-One (AB1; 4×4 grid).

● Each slice is partitioned into a 4×4 grid (16 patches).
● For each patch position, that patch is masked as the target region, and the remaining patches provide context as input.
● Up to 16 pairs per slice are generated (one per target patch).

Symmetry (left–right hemisphere prediction).

● Each slice is divided into left and right halves.
● One half is used as input, and the contralateral half is the target region.

Empty-target exclusion. Pairs were excluded if the target region contained no brain signal after preprocessing (i.e., target region is empty) to avoid training on non-informative background.

Model input. For both strategies, the reconstructor input is a two-channel tensor formed by concatenating:

1. the masked intensity slice, and
2. the aligned edge-map channel.

Pair counts (BrainMetShare Mets normal corpus).

From 9,772 normal slices, AB1 yields 156,352 pairs, and Symmetry yields 19,544 pairs.

Augmentation. Standard 2D augmentations (e.g., shear and horizontal flipping) were applied to masked pairs, consistently across channels and targets.

### S1.3 Reconstruction model and optimization

Architecture. A 2D U-Net reconstructor (input 256×256×2) was trained for each configuration. Each model has 23,602,177 parameters.

Training configurations. Four reconstruction variants were trained:

● AB1 + MSE
● AB1 + ROI-restricted loss
● Symmetry + MSE
● Symmetry + ROI-restricted loss

Loss functions.

● **MSE**: build-in loss metric
● MSE: build-in loss metric

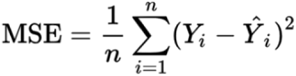

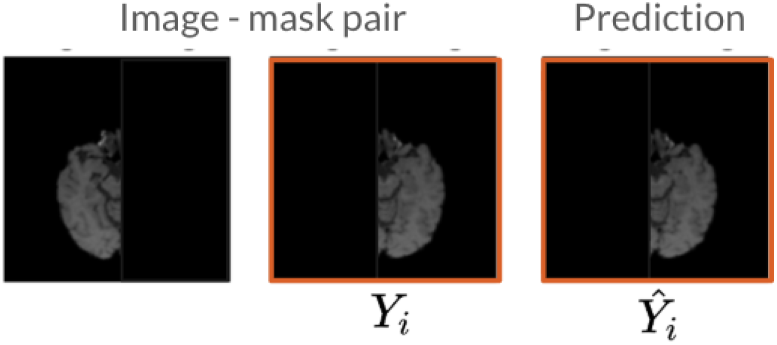

● ROI loss: customize loss only focus within brain area using contour map

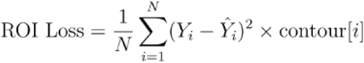

Where:

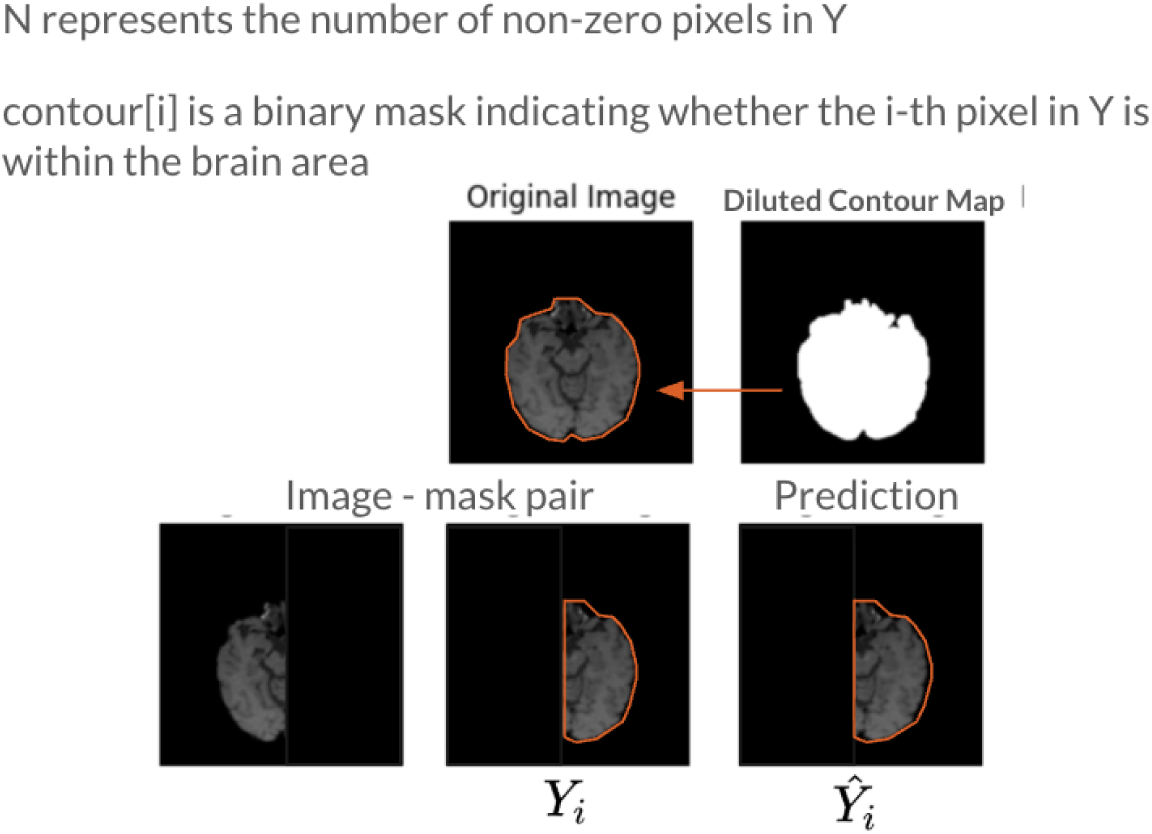

Training schedule and checkpointing.

● Maximum training budget: 300 epochs
● Best checkpoint selected by lowest validation forward-pass loss (reported in Table 2 in the main text).

Reproducibility parameters (fill from run configs):

● Optimizer = [TBD]
● Learning rate and schedule = [TBD]
● Batch size = [TBD]
● Weight decay / regularization = [TBD]
● Checkpoint metric definition (exact tensor/region) = [TBD]

### S1.4 Difference representation and Beta-VAE embeddings

After U-Net training, each slice was reconstructed to obtain a predicted disorder-free slice, and a difference map was computed between the original slice and its reconstruction.

Beta-VAE training.

● Input: per-slice difference maps
● Output: 1024-dimensional latent embedding per slice
● Separate Beta-VAE models were trained for each reconstruction configuration (masking strategy × loss).

Reproducibility parameters (fill from run configs):

● Beta-VAE architecture details = [TBD]
● β (KL weight) and any annealing schedule = [TBD]
● Optimizer/LR/batch size/epochs = [TBD]

## Appendix S2. Additional evaluation protocols and sensitivity analyses

**Table S1.**
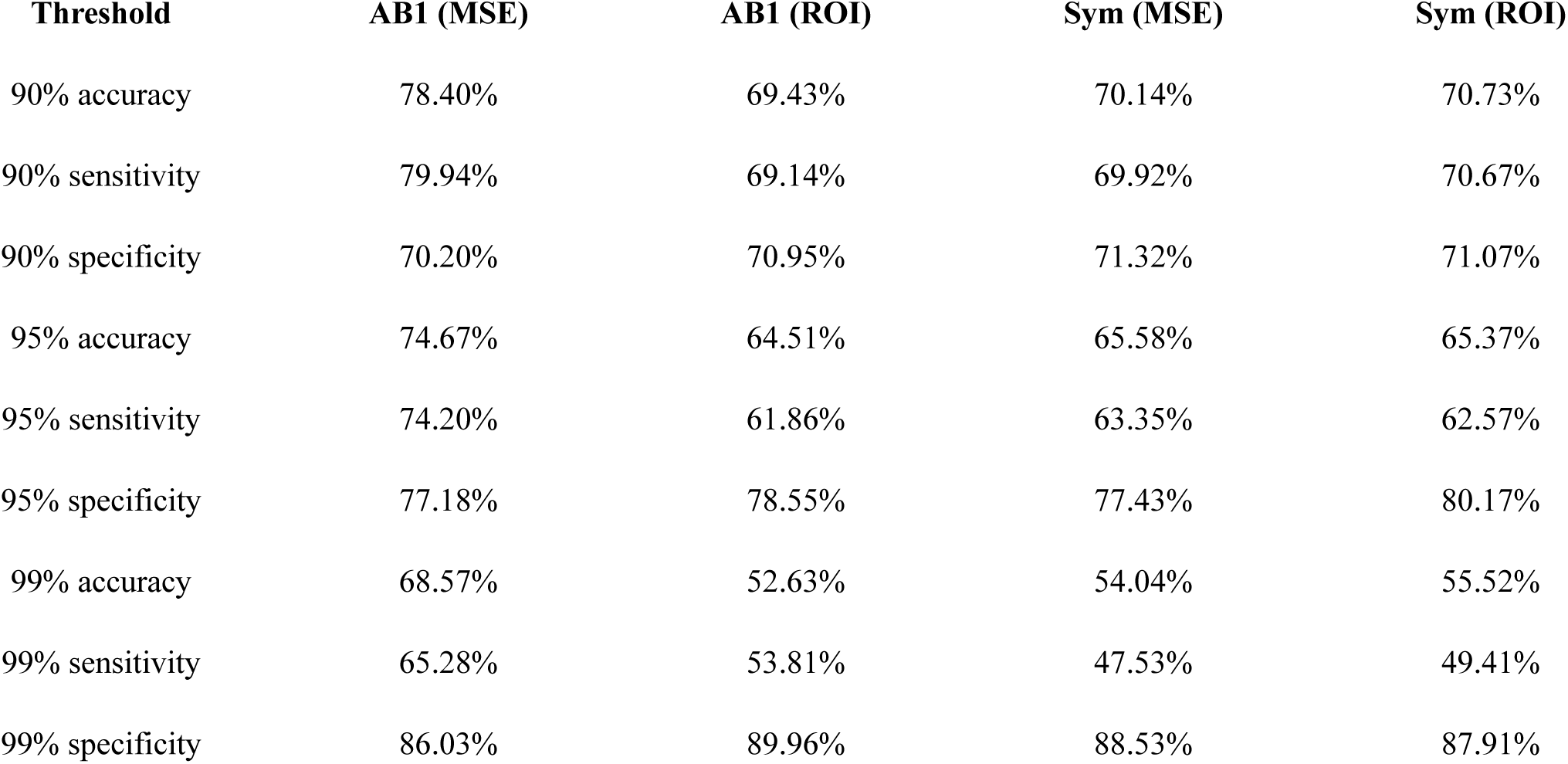
Mets anomaly detection accuracy with half validation-normal as reference.

### S2.3 External adaptation under domain shift: fine-tuning protocol and ROC thresholding

For SSA and Alz, we fine-tuned the pretrained U-Net reconstructor for 50 epochs using normal slices only, while keeping the Beta-VAE frozen (no gradient updates). We varied the amount of available normal training data: 0%, 25%, 50%, 100% (0% denotes no fine-tuning).

Threshold selection (ROC-optimized). Operating thresholds were selected on the validation split by maximizing Youden’s J statistic: TPR - FPR, using validation-normal and labeled abnormal slices. Performance was then reported on the test split at the selected threshold using accuracy, sensitivity, and specificity.

**Table S2.**
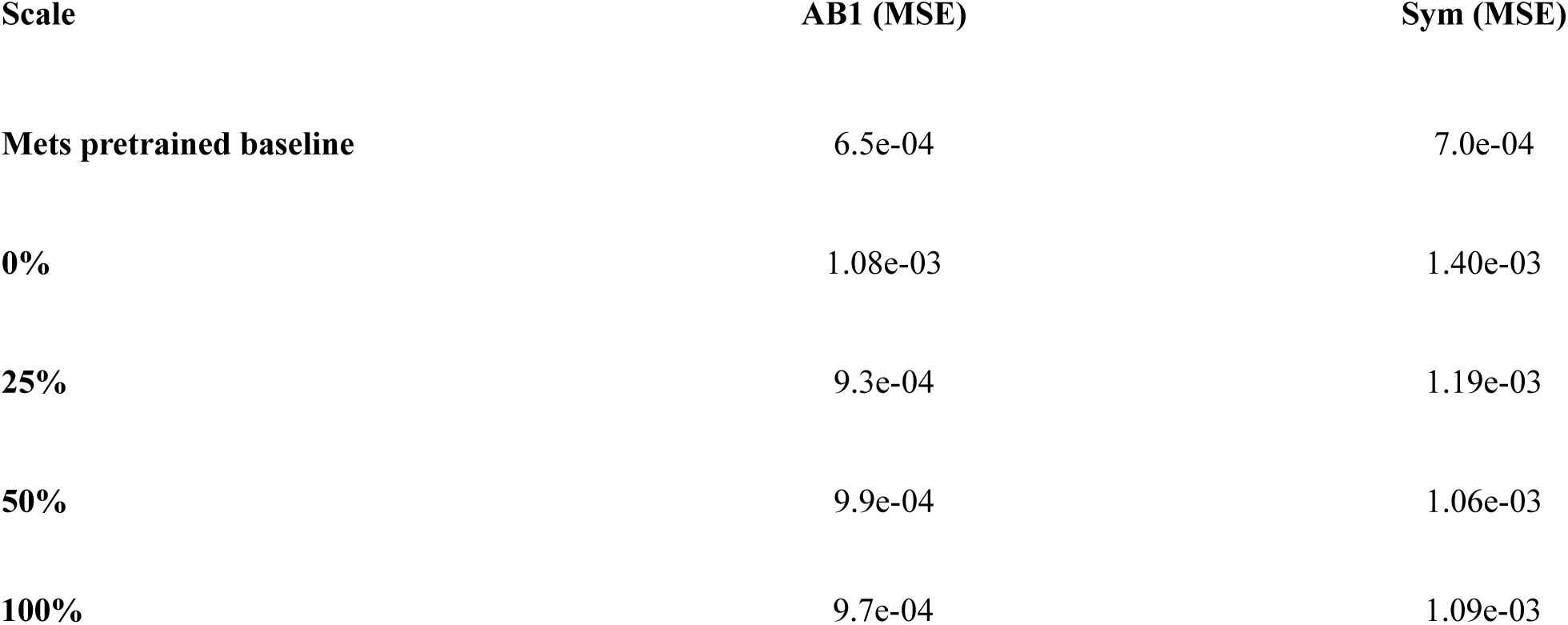
SSA reconstruction loss after fine-tuning (MSE models).

**Table S3.** Alz reconstruction loss after fine-tuning.

| Scale | AB1 (MSE) | Sym (MSE) |
| --- | --- | --- |
| Mets pretrained baseline | 6.5e-04 | 7.0e-04 |
| 0% | 7.36e-03 | 1.24e-02 |
| 25% | 3.48e-04 | 4.86e-03 |
| 50% | 3.31e-04 | 4.54e-03 |
| 100% | 3.28e-04 | 4.36e-03 |

**Table S4.** Alzheimer’s (Alz) anomaly classification under domain shift using ROC-optimized Mahalanobis thresholding across fine-tuning scales.

| Threshold |  | Finetune Scale |  |  |  |
| --- | --- | --- | --- | --- | --- |
|  |  | 0% | 25% | 50% | 100% |
| Ab1 ROC | Accuracy | 65.12% | 65.12% | 61.19% | 70.54% |
|  | Sensitivity | 66.16% | 65.81% | 61.62% | 71.84% |
|  | Specificity | 44.38% | 51.25% | 52.5% | 44.38% |
| Sym ROC | Accuracy | 36.99% | 51.07% | 49.67% | 43.96% |
|  | Sensitivity | 34.66% | 50.28% | 48.56% | 42.96% |
|  | Specificity | 83.75% | 66.88% | 71.88% | 73.12% |

